# Differentiating Hyperkinetic and Hypokinetic Motor Features in the Progression of Huntington’s Disease

**DOI:** 10.1101/2025.04.17.25325819

**Authors:** Nabil Halabi, Annie Killoran, Peg C. Nopoulos, Jordan L. Schultz

## Abstract

**Background:** Huntington’s disease (HD) is a monogenic neurodegenerative disorder typically characterized by chorea, a hyperkinetic motor feature. Historical data suggest that hypokinetic features, like rigidity and bradykinesia, become more prominent in later stages of HD. No evidence-based analysis has confirmed this observation. Additionally, several motor features of the disease are not clearly defined as hypokinetic or hyperkinetic.

**Objectives:** This study aimed to 1) elucidate the trajectory of hyperkinetic and hypokinetic features across the disease course and 2) to classify vague motor features as following a hyperkinetic or hypokinetic trajectory.

**Methods:** Data from 13,475 motor-manifest HD patients from the Enroll-HD platform were analyzed. Linear mixed-effects models were constructed for each of the 31 Unified Huntington’s Disease Rating Scale (UHDRS) motor subscales, with disease burden as the primary predictor. The models were used to generate the trajectories of features known to represent hyperkinesis and hypokinesis, with the same being done for vague subscales. Dynamic time warping (DTW) was then used to classify said subscales as having a hyperkinetic or hypokinetic trajectory.

**Results:** Hyperkinetic features rise initially and diminish in middle disease, while hypokinetic features continually increase across the disease course. All non-choreiform features demonstrated a hypokinetic-like trajectory.

**Conclusions:** HD is generally considered a hyperkinetic movement disorder, but the middle and late stages of the disease are predominated by hypokinesis. These findings suggest that hypokinetic features may be a larger contributor to the overall motor burden of HD. This has significant implications for clinical trial design, motor phenotype clustering, and pharmacotherapy.

## 1. Introduction

Huntington’s disease (HD) is the most-common monogenic neurodegenerative disorder worldwide(1), as well as the most-common inherited form of dementia(1). The disorder is caused by a trinucleotide repeat expansion of cytosine-adenine-guanine (CAG) in the huntingtin (*HTT*) gene, found on chromosome 4p16.3(2). The symptomatology is multifaceted, including characteristic motor, psychiatric and cognitive manifestations (3). The age of motor onset varies based on the number of CAG repeats but typically occurs during the fourth to sixth decades of life(4). In cases of CAG repeats upwards of 55 copies, patients may develop symptoms during their childhood/adolescence, referred to as juvenile-onset HD (JoHD).(5)

Chorea is a defining feature of HD. These hyperkinetic movements can be quite conspicuous, but many patients lack insight into their chorea. Historical data has shown that these movements wane in the later stages of the disease(6), and hypokinetic features, such as rigidity and bradykinesia, become more pronounced.(7) While clinicians have observed this through clinical practice, no data-based analysis has demonstrated the trajectory of hyperkinetic or hypokinetic motor features across the disease course.

Studies have shown that patients with more hypokinetic features such as rigidity and bradykinesia tend to be globally worse and have lower total functional capacity (i.e. TFC).(8) Notably, bradykinesia strongly influences gait disturbance, which may contribute to an increased risk for falls and subsequent injury, further limiting functionality. (9) Furthermore, akinetic features have also been shown to be inversely related to measures of cognitive performance, further illustrating the fidelity of hypokinetic or parkinsonian features in tracking the disease course.(10) Conversely, studies have demonstrated a *lack* of relationship between hyperkinetic features and cognitive manifestations.(10) Additionally, a recent investigation into the motor phenotypes of HD described patients with a predominance of chorea as having (on average): less CAG repeats, larger pallidal volumes, shorter disease duration, larger whole-brain volume and a more benign course of psychiatric symptoms.(11) Therefore understanding the trajectory and relative contributions of hyperkinetic and hypokinetic features could provide important insights into both how these subscales change over the disease course, and the impact they may have on burden and functional capacity for patients.

Currently, the Unified Huntington’s Disease Rating Scale Total Motor Score (UHDRS-TMS) is the gold standard for measuring the burden of motor features in HD.(12) The TMS is comprised of 31 subscales that includes measures of hypokinesis, such as bradykinesia, and hyperkinesis, such as chorea. However, there are several subscales that are not as clearly distinguishable, such as dysarthria or ocular saccades(13). For example, dysarthria was classically considered a hyperkinetic feature(14), but recent evidence has challenged this notion(15). Several UHDRS items(16) occupy a-vague territory with uncertain pathoetiology. It has yet to be demonstrated whether these undefined features follow a more hyperkinetic or hypokinetic trajectory across the disease course. This information may have profound implications related to motor phenotype clustering, clinical trial design, and tailored pharmacotherapy for patients.

Utilizing the Enroll-HD database, the current study is designed to evaluate the relative severity of hyperkinetic and hypokinetic motor features across the HD disease course. In addition, features that are not considered to be canonically hyperkinetic or hypokinetic will be evaluated to determine if their course over time can be classified as one or the other based on trajectory.

## Methods

### 1.1. Study Design

We leveraged data from the *Enroll-HD* platform, including data from subjects previously followed in the predecessor *REGISTRY* European study.(17) *Enroll-HD* is an international observational study that periodically records information from HD patients (and controls) approximately every year. (17) *Enroll-HD* has thousands of participants, with motor evaluations (i.e. UHDRS-TMS) performed at each study visit. This analysis included data from subjects with only one available visit and data from subjects with more than one visit. Utilizing all available data leads to better representation across the disease spectrum to more accurately model the trajectory of motor subscales, similar to accelerated longitudinal design(18).This design allows UHDRS motor symptoms to be modeled over time based on disease burden (DB = [CAG-35.5] X age), creating a trajectory of severity of symptoms with increasing DB scores representing disease progression (19).

A higher number of CAG repeats is associated with faster progression of motor features (19). Disease burden is used to account for the differential change in subscale severities over time as a function of a person’s age and CAG repeat length. Milestones of the disease occur at fairly predictable disease burden ranges: pre-manifest HD begins roughly around a burden of 250(20), achieving motor conversion when the burden reaches the 300 range(21), middle disease at a range of 400(22), and fewer, longer-lived patients experiencing burdens in the 800-1000 range(23). As such, only visits where participants’ disease burden was between 200 to 800 were used, to properly represent the average course of motor features. After the exclusion criteria were applied, the total number of participants included for analysis was 13,475, constituting 49,305 visits (*Figure 1*).

**Figure 1:**
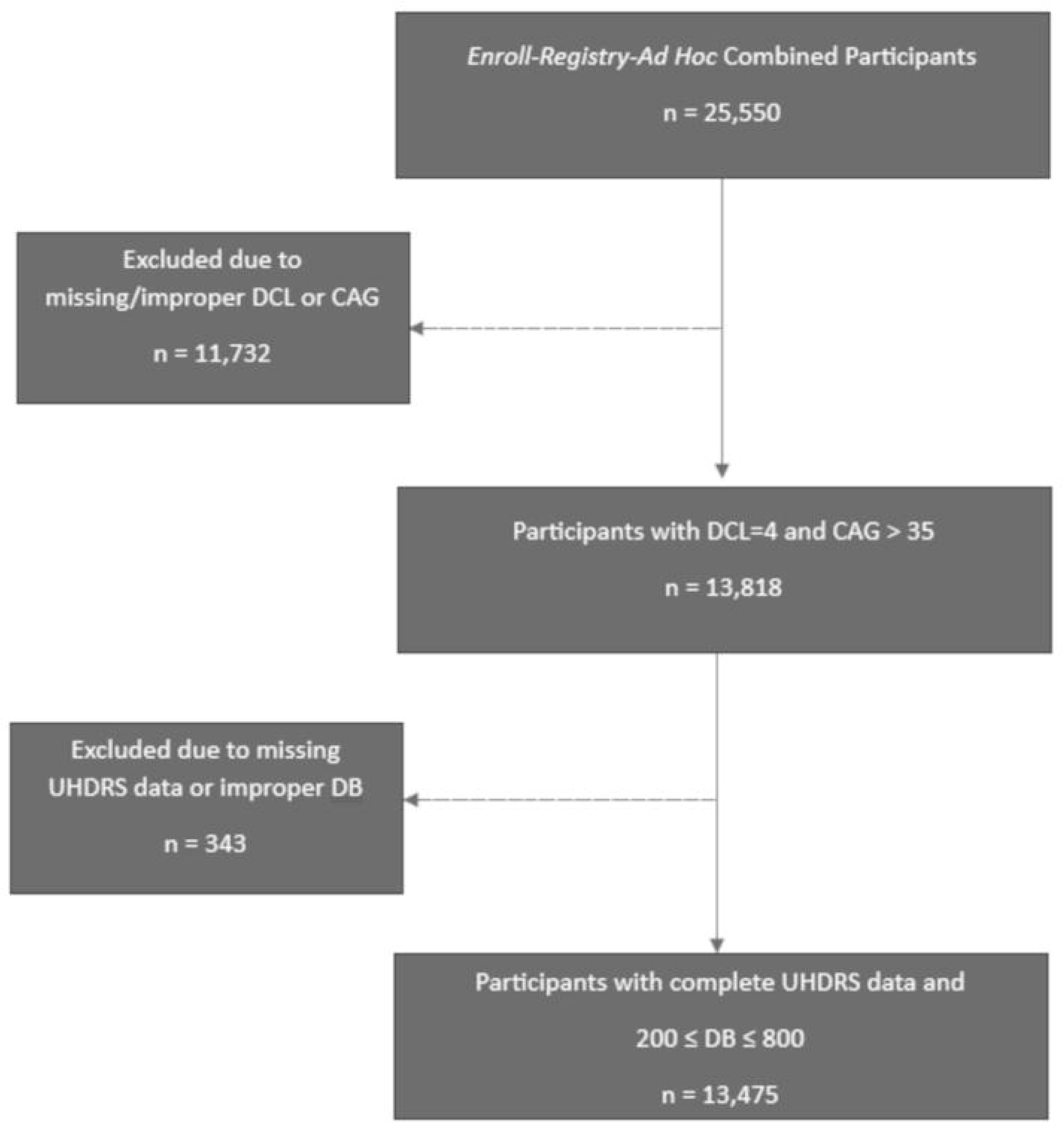
“Flowchart of Inclusion Criteria.” Abbreviations: DCL, diagnostic confidence level; CAG, cytosine-adenine-guanine; UHDRS, united Huntington’s Disease rating scale; DB, disease burden.

Due to our focus on motor feature progression, analysis was limited to motor-manifest HD patients who had a diagnostic confidence level (DCL) of 4 (indicating 99% diagnostic confidence in pronouncing HD) and a CAG repeat length greater than 35. Controls were therefore filtered out of analysis, in addition to those with missing diagnostic information (i.e. an unknown DCL). For participants who met the initial criteria, their UHDRS motor scores for all 31 items across each study visit were extracted. Participant visits with missing datapoints for any of the UHDRS motor items were excluded from further analysis.

### 1.2. Statistical Analysis

Statistical analysis was performed using the *R* programming language (version 4.4.1) in *Rstudio* 2023.03.1 Build 446. We constructed linear mixed-effects models for UHDRS hyperkinetic and hypokinetic subscales using the severity scores and disease burdens measured at each study visit. Each model specified severity of the motor feature (ranging from 0-4 in whole number increments) as the dependent variable. The independent variable was disease burden, to which a natural spline transformation with 3 degrees of freedom was applied, allowing for non-linear relationships to be evaluated. We included assigned gender at birth, antipsychotic use, and vesicular monoamine transporter type 2 inhibitor (VMAT2i) use as fixed-effect predictors in the models. Disease burden accounts for age and CAG, so these individual measures were not included to avoid co-linearity. Subject ID (a unique identifier for each participant) was included as a random-effect predictor, accounting for intersubject and intrasubject variability.

To verify the utility of the burden term in capturing disease course, an analysis of variance (i.e. ANOVA) was performed. This involved comparing models with and without the non-linear ‘burden’ predictor. A significant result from the ANOVA indicates that the inclusion of ‘burden’ significantly captures variation across disease course. After validating the linear mixed-effects models, we generated predicted severities of hyperkinetic and hypokinetic motor features across disease burdens 200 to 800.

To address our first aim, we created averaged subscale trajectories for canonical hyperkinetic and hypokinetic features. These averaged trajectories were constructed by taking the mean values of the predicted severities from the hyperkinetic and hypokinetic subscales. The representative hyperkinetic motor features included the chorea subitems [facial, buccal-oral-lingual (BOL), truncal, right upper extremity (RUE), left upper extremity (LUE), right lower extremity (RLE) and left lower extremity (LLE)]. Similarly, the representative hypokinetic trajectory was composed by averaging the values of bradykinesia, right-arm rigidity, left-arm rigidity and retropulsion pull test (24). Once the averaged values were calculated, they were plotted by disease burden, generating summarized trajectories for hyperkinesis and hypokinesis.

For the second aim, equivalent linear models were constructed and predicted severity values were generated for the remaining 20 ‘vague’ motor subscales. A statistical method for classification was employed which rests on the assumption that similarity in disease course trajectory implies similarity in pathoetiology. This assumption is based on indirect causality and may not accurately represent neuropathology. Nevertheless, it remains a useful surrogate measure (in the absence of definitive evidence) for inference into the nature of motor features.

The method in which motor features were categorized as either hyper-or hypokinetic was through ‘dynamic time warping’ (DTW). DTW is a type of time series analysis that assesses the degree of similarity between two curves. Like the calculation of Euclidean distance between two curves, DTW aligns datapoints in one series to points in its comparator. Where Euclidean distance and DTW differ, however, is the alignment process. When two series are misaligned, it is inaccurate to align datapoints in a one-to-one fashion. Instead, the DTW algorithm aligns datapoints in a one-to-many and many-to-one fashion, to preserve as much accuracy as possible.

After inputting a given curve, the DTW function outputs the summative distance between the inputted curve and a standard reference curve. For the second aim, the standard reference curves were the averaged hyperkinetic and hypokinetic trajectories. These distances from each standard reference curve for each of the 20 remaining motor subscales were compared, and the subscale was classified as hyperkinetic or hypokinetic based on the lower DTW distance.

Bootstrap resampling was performed to assess the robustness of this classification, with 100 iterations of resampling conducted for each subscale. For each iteration, the resampled data was used to refit models for each subscale and generate predicted curves to be input into the DTW function. The average DTW distance difference (i.e. distance to hyperkinetic minus distance to hypokinetic) was calculated. The confidence intervals of these differences were derived from the bootstrap iterations, and features were classified as hyperkinetic or hypokinetic based on the mean difference in distances. If the mean difference was above zero and the confidence interval did not cross zero, the subscale was classified as hyperkinetic. Conversely, if the mean difference was below zero and the confidence interval did not cross zero, the subscale was classified as hypokinetic.

### 1.3. Ethical Considerations

The *Enroll-HD* clinical platform, funded and run by CHDI (Cure Huntington’s Disease Foundation), has conducted their original study under the oversight and approval of multiple Institutional Review Boards. More specifically, each *Enroll-HD* and *Registry* clinical site has their own consent form and consent-obtaining process, overseen by the local IRB at their institution. Part of this consent involves informing participants that their *de-identified* data may be made available to qualified researchers at a later date.

Our research team at the University of Iowa has signed a data-sharing agreement between ourselves and *Enroll-HD* that specifies the sharing of *de-identified* participant data from *Enroll-HD.* Finally, our study was reviewed by our Institutional Review Board (i.e. at the University of Iowa) and determined to be a secondary data analysis of the *Enroll-HD* data base and not human subject’s research requiring IRB approval; however, the Chair of the IRB Committee provided confirmation that it was ethical for us to conduct this retrospective research.

### 1.4. Data Sharing

The datasets utilized for this secondary analysis are obtainable from the *Enroll-HD* clinical research platform, courtesy of CHDI, for use by professional researchers in their studies on HD.

## Results

This study included 13,475 subjects (*Table 1*). Age was distributed across a large range, spanning from 9 to 93 years of age, with a mean age of 53.3 ± 12.7 years-old. The division between people assigned male at birth (AMAB) and assigned female at birth (AFAB) was roughly equal, being split 48.1% (6,481) to 51.9% (6,994), respectively. Approximately half (48.4%) of all participants used antipsychotics, while only a quarter (25.1%) reported using VMAT2i’s. Almost three-fifths (58.8%) reported using any form of antidopaminergic therapy. The average disease burden at baseline was 419 ± 91.3, ranging between the predefined limits of 200 to 800. The UHDRS-TMS has a maximum value of 124, derived by summating all 31 testable items. The average TMS at baseline in this cohort was 37.9 ± 19.5, with values ranging from 0 to 116. The cohort was predominantly Caucasian (93.5%), which is concordant with literature citing a predominance of HD in White populations.(25) Alternatively, this could be interpreted as selection bias due to *Enroll-HD* sites being located in majority-White locations.(17) Finally, the average CAG repeat length was 44.0 ± 3.77, with values ranging from 38 to 70.

**Table 1:** Baseline Demographics of Study Cohort.

| Demographic | Value |
| --- | --- |
| **Age (yrs.)** |  |
| Mean (SD) | 53.3 (12.7) |
| Median [Min, Max] | 53.7 [9.34, 93.0] |
| **Assigned Gender at Birth** |  |
| Assigned Male at Birth (%) | 6,481 (48.1%) |
| Assigned Female at Birth (%) | 6,994 (51.9%) |
| **Calculated BMI (kg/m<sup>2</sup>)** |  |
| Mean (SD) | 24.9 (4.86) |
| Median [Min, Max] | 24.2 [11.7, 64.7] |
| Missing (%) | 289 (2.1%) |
| **Duration of Disease (yrs.)** |  |
| Mean (SD) | 6.10 (4.78) |
| Median [Min, Max] | 5.00 [0, 45.0] |
| Missing | 569 (4.2%) |
| **Age at Clinical Diagnosis** |  |
| Mean (SD) | 49.0 (12.6) |
| Median [Min, Max] | 49.0 [4.00, 92.0] |
| Missing (%) | 569 (4.2%) |
| **Use of Antipsychotics** |  |
| Users (%) | 6,522 (48.4%) |
| Never-Users (%) | 6,953 (51.6%) |
| **Use of VMAT2 Inhibitors** |  |
| Users (%) | 3,385 (25.1%) |
| Never-Users (%) | 10,090 (74.9%) |
| **Use of Antidopaminergic Medications** |  |
| Users (%) | 7,921 (58.8%) |
| Never-Users (%) | 5,554 (41.2%) |
| **Disease Burden** |  |
| Mean (SD) | 416 (91.3) |
| Median [Min, Max] | 410 [200, 800] |
| **Age of Death (yrs.)** |  |
| Mean (SD) | 60.8 (12.6) |
| Median [Min, Max] | 61.0 [27.0, 95.0] |
| Missing (%) | 11,992 (89.0%) |
| **Cause of Death** |  |
| Pneumonia (%) | 251 (1.9%) |
| Other Infection (%) | 70 (0.5%) |
| Cancer (%) | 50 (0.4%) |
| Stroke (%) | 26 (0.2%) |
| Trauma (%) | 38 (0.3%) |
| Suicide (%) | 72 (0.5%) |
| Other (%) | 520 (3.9%) |
| Missing (%) | 12,448 (92.4%) |
| **Total Motor Score** |  |
| Mean (SD) | 37.9 (19.5) |
| Median [Min, Max] | 35.3 [0, 116] |
| **Ethnicity** |  |
| White (%) | 12,597 (93.5%) |
| Black (%) | 104 (0.8%) |
| Latinx (%) | 330 (2.4%) |
| Native American (%) | 42 (0.3%) |
| Mixed (%) | 104 (0.8%) |
| Asian (%) | 86 (0.6%) |
| Other (%) | 207 (1.5%) |
| Missing (%) | 5 (0.0%) |
| **CAG Repeats** |  |
| Mean (SD) | 44.0 (3.77) |
| Median [Min, Max] | 43.0 [38.0, 70.0] |

Abbreviations: Yrs, years; BMI, body mass index; kg, kilogram; m<sup>2</sup>, squared meter; SD, standard deviation; VMAT2, vesicular monoamine transporter type-2; CAG, cytosine-adenine-guanine; min, minimum; max, maximum.

Visually, the averaged trajectories of the representative hyperkinetic and hypokinetic curves differ (*Figure 2*). The hyperkinetic trajectory curve resembles a negative parabola, demonstrating a rise and fall in severity, with a vertex (maximum severity) between 1 and 2 at a disease burden of roughly 500. This is significant because it demonstrates that chorea (a hallmark of the disease) is quantified as quite mild, peaking at an average rating of only 1.5 units before diminishing at mid-disease. This is contrasted by the hypokinetic trajectory curve, which resembles a parabolic segment with strictly positive inflection, increasing indefinitely to an observed maximum severity between 3 and 4 at a burden of 800.

**Figure 2:**
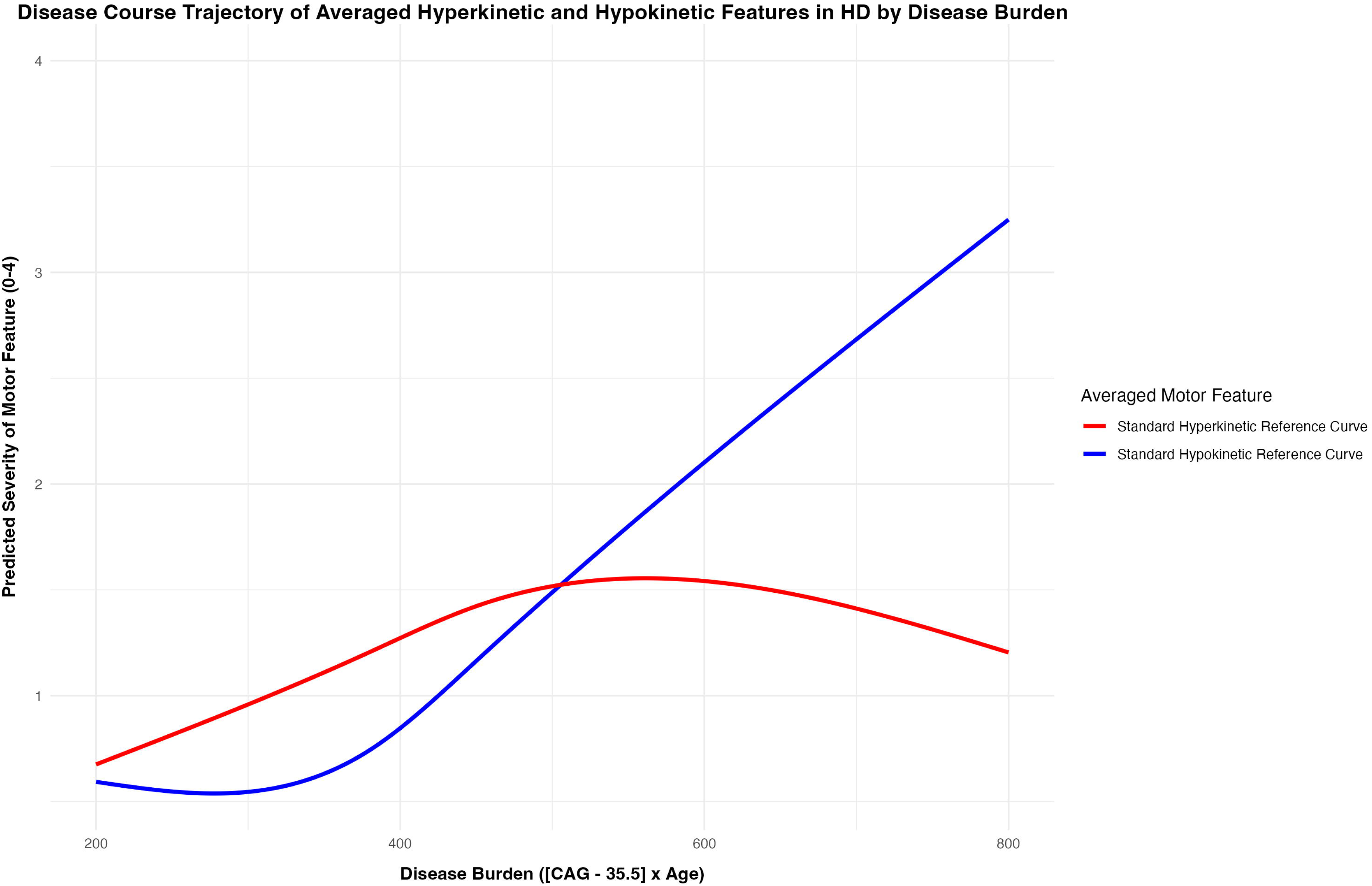
“Disease Course Trajectory of Averaged Hyperkinetic and Hypokinetic Features in HD by Disease Burden.” Abbreviation: CAG, cytosine-adenine-guanine; HD, Huntington’s Disease.

The second aim categorized the remaining UHDRS motor subscales that were not classically defined as either hyper-or hypokinetic. By comparing each subscales’ disease course trajectory to both the hyper– and hypokinetic trajectories (figure 2), we classified each motor feature depending upon how closely they aligned with either trajectory. The DTW analyses classified the 20 remaining subscales as hypokinetic (*Supplementary Table 2*). Specifically, gait (DTW_hypo_ = 55.7 versus DTW_hyper_ = 599.6), finger-tapping left (DTW_hypo_ = 132.5 versus DTW_hyper_ = 770.5) and saccade velocity horizontal (DTW_hypo_ = 69.3 versus DTW_hyper_ = 630.6) were most closely related to the hypokinetic trajectory. However, this sharp distinction in classification was not unanimous amongst all subscales. Notably, dystonia of the right lower (DTW_hypo_ = 177.9 versus DTW_hyper_ = 230.3) and left lower extremities (DTW_hypo_ = 181.2 versus DTW_hyper_ = 231.2) displayed the least separation between the two trajectories. Dystonia of the trunk (DTW_hypo_ = 79.0 versus DTW_hyper_ = 315.0) and the right upper (DTW_hypo_ = 88.3 versus DTW_hyper_ = 315.8) and left upper extremities (DTW_hypo_ = 91.9 versus DTW_hyper_ = 315.1) also share in this trend. This agrees with evidence on the ambiguity of classifying dystonia as hyper– or hypokinetic.(26) In addition, the classification of features using bootstrapping agrees with the simplistic DTW classification. The bootstrapped results can be found in *Table 2* and *Figure 3*.

**Figure 3:**
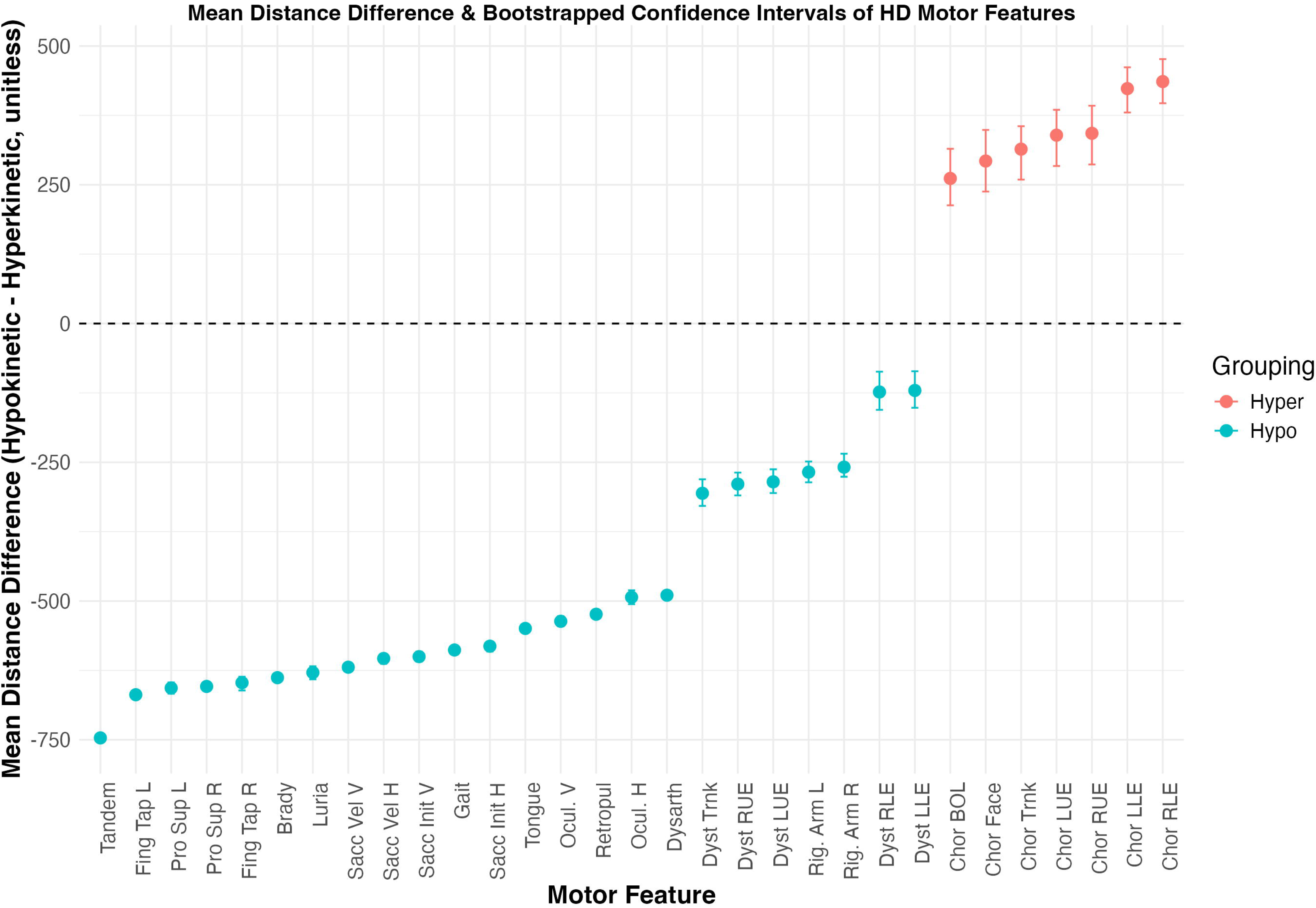
“Mean Distance Difference & Bootstrapped Confidence Intervals of HD Motor Features.” Abbreviations: UHDRS, united Huntington’s Disease rating scale; R, right; L, left; UE, upper extremity; LE, lower extremity; chor, chorea; dyst, dystonia; vel, velocity; sacc, saccadic; ocul, ocular; fing tap, finger tapping; pro sup, pronation supination; brady, bradykinesia, retropul, retropulsion test.

**Table 2:**
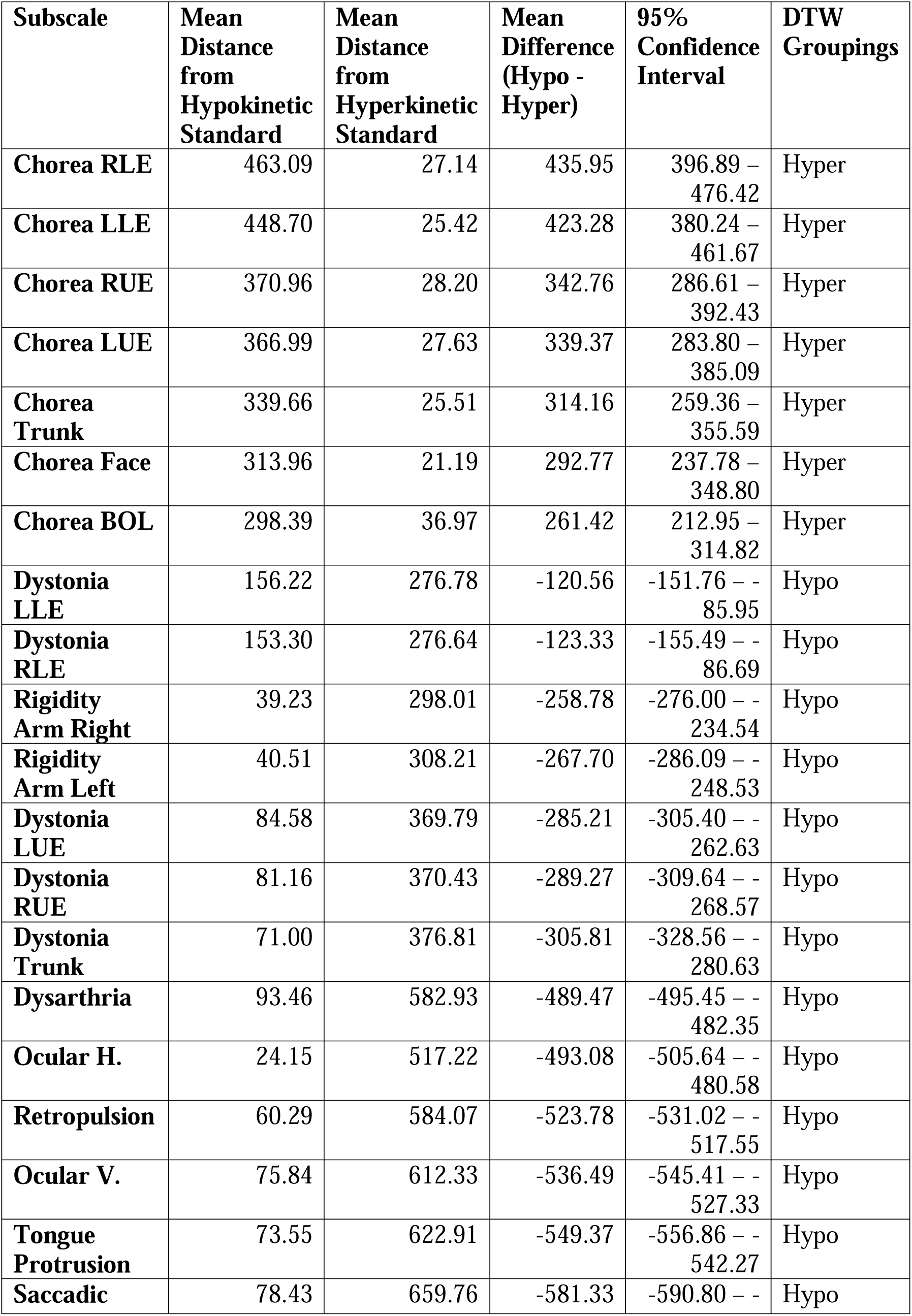
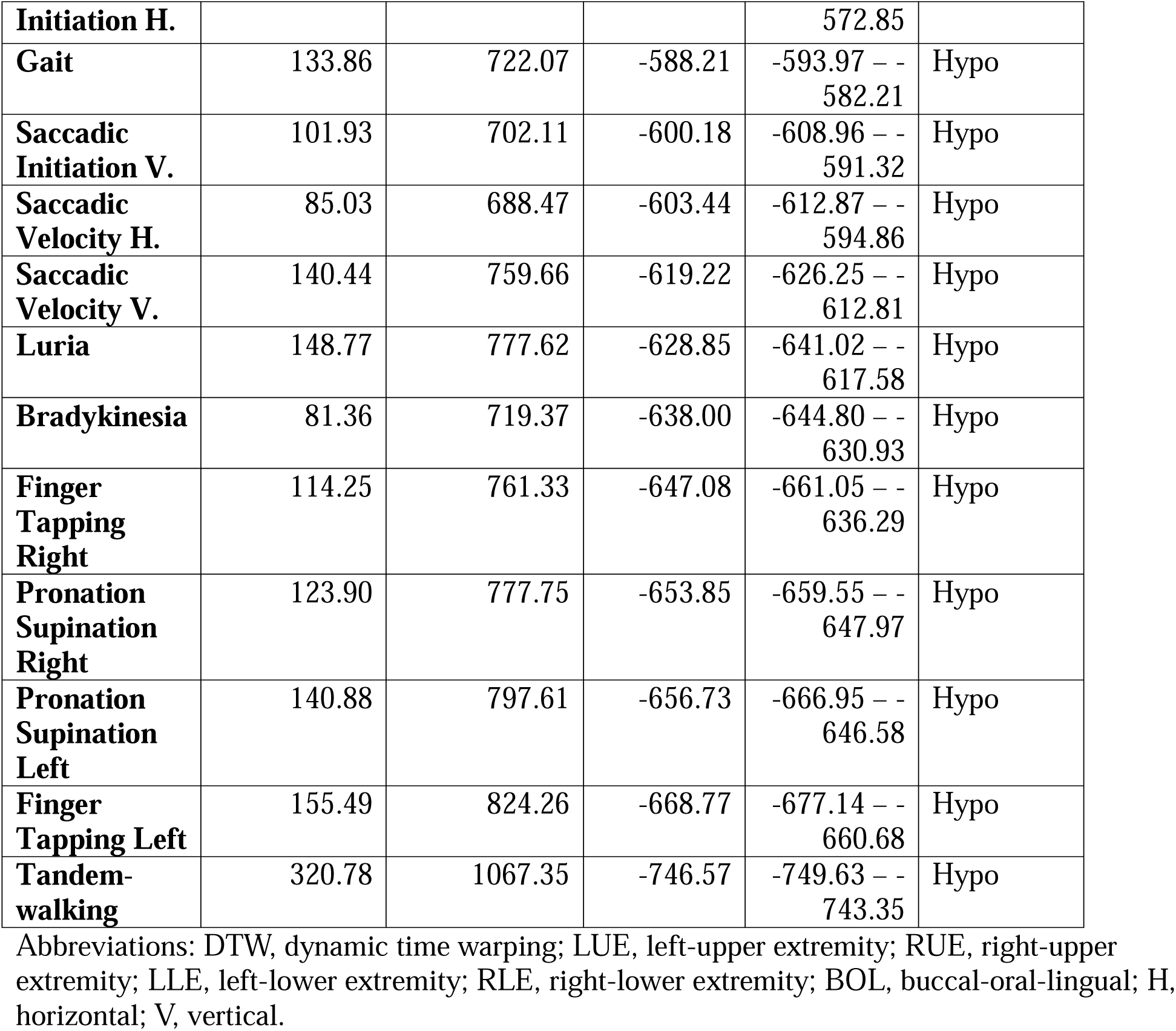
Motor Subscale Classification by Mean Distance Difference and Bootstrapped Confidence Intervals.

## Discussion

Our findings show that all hyperkinetic and hypokinetic subscales change as a function of disease burden, but the pattern of change between hypokinetic and hyperkinetic features varied widely. This emphasizes the dynamic nature of the motor phenotype in HD. Specifically, chorea subscales seemed to be the predominant motor feature early in the disease; however, chorea plateaued at a relatively low severity (on average) in the mid-range of the disease before beginning to decrease. In contrast, hypokinetic features emerged later in the disease but demonstrated more profound change with time and no plateau or upper limit on the severity. This is significant because it demonstrates that as disease burden increases, the proportion and magnitude of hypokinetic features is more likely to continually increase. These findings re-evaluate historical data describing the progression of motor features in HD, where hypokinesis is typically thought to occur only in late disease.(27) By contrast, our findings demonstrate that hypokinesis becomes pre-eminent in the middle of the disease. Furthermore, our findings that hypokinetic features display more significant and continuous change across disease course when compared to hyperkinetic ones is similar to a previous report (16) where motor feature progression in JoHD was investigated and showed that hypokinetic features progress in a more consistent and predictable fashion across disease course than hyperkinetic ones. Although the rate of TMS progression in JOHD is faster than that of AoHD, the findings that are consistent in both reports are that the hypokinetic features are the most prominent motor feature with continuous progression over time, sharing the same trajectory over disease course.

Paired with observations that hypokinetic features track cognitive and functional measures, this provides compelling evidence that hypokinesis is a significant part of HD pathology. The inflection point at disease burden 400-500 demonstrates the simultaneous presence of mixed motor features during middle disease. In summary, our findings demonstrate that although hyperkinesis and hypokinesis co-exist simultaneously, much of the motor burden of HD may be due to hypokinetic features, especially in the mid to late stages of the disease. This is analogous to the presence of mixed features in other movement disorders classically considered monomorphic (e.g. the presence of (hyperkinetic) tremor in Parkinson’s Disease, which is classified as a bradykinetic disorder).

The secondary findings show that all non-choreiform subscales of the UHDRS-TMS follow a trajectory more akin to hypokinesis. This finding is interesting given that chorea has been considered the most prominent and distinguishing feature of HD.(28), presumably because it is so visually apparent. As such, it was expected that motor abnormalities would exist across a spectrum; with some features progressing similar to hyperkinesis, while others would follow a hypokinetic trajectory.(29) Rather, these findings indicate that all non-choreiform features progress similarly to bradykinesia and rigidity. While these features were significantly classified as hypokinetic using DTW, the degree of similarity to hypokinetic progression did vary. For example, dystonia in the left and right lower extremities showed a mean distance difference of – 120.56 and –123.33 units respectively (*Table 2*). In comparison, the mean difference for tandem walking was –746.57 units (*Table 2*). These differences indicate that while there are clear distinctions between the hyperkinetic and the hypokinetic features, ‘hypokinetic’ features still display heterogeneity with regards to their trajectory.

These findings may have significant clinical implications. First, these findings may help to optimize the timing of antichorea treatments, such as 1^st^ and 2^nd^ generation antipsychotics and VMAT2i’s (tetrabenazine, deutetrabenazine and valbenazine). While these antidopaminergics are effective treatment options for chorea (30), they have potential adverse effects, such as drug-induced parkinsonism (7, 9) (31). Anecdotally, antidopaminergic agents are often discontinued in patients with HD when chorea becomes noticeably reduced, but generally this does not occur until patients reach later stages of the disease. As such, the use of antidopaminergic medications may have a more negative impact on hypokinetic features earlier in the disease course of HD. This sentiment was outlined in the reporting of results from the TETRA-HD study (30). There, the authors provided “(a) reminder that chorea is only one of many sources of disability for patients with Huntington’s disease.”(30) While only 4% of participants receiving tetrabenazine in the TETA-HD study experienced drug-induced parkinsonism, they did find that tetrabenazine significantly lowered functional scores, as TFC was positively correlated with worsening measures of parkinsonism.(30) This is not to say that the use of antidopaminergic medications in HD is improper. Rather, these findings highlight that the global UHDRS-TMS score maybe an insufficient tool when considering antichorea treatment for patients with HD. Instead, clinicians should carefully consider both their patients’ hypokinetic and hyperkinetic findings within the UHDRS-TMS when making such treatment decisions. Furthermore, monitoring should be done on an ongoing basis as the hypo– and hyperkinetic features evolve with disease progression.

This study benefits from many participants (over 10,000) and an accelerated longitudinal design, instilling it with the necessary power to distinguish true trends from chance. Furthermore, data on multiple covariates was incorporated into the statistical modelling, providing defense against spurious correlations. The validity of said models was verified by ANOVA, and cluster assignments were triangulated by various methods, including dynamic time warping and bootstrap resampling. This provides considerable weight to our claims that hypokinesis predominates the landscape of motor features in HD.

There are some aspects of the study design which have limitations, however. Firstly, since the UHDRS is administered by an examiner,(12) there is room for intra-examiner and inter-examiner variability in the assignment of severity levels. The descriptions of motor abnormalities by the UHDRS are not always well-defined and could be open to interpretation.(12) Additionally, the inter-visit interval of 1-year may not capture the subtle variations in feature severity that a tighter follow-up window would have. The limited number of JoHD participants may also limit the study’s generalizability to all variants of HD, despite similar results obtained with respect to the trajectory of motor features in AoHD and JoHD(16). Although antipsychotic and VMAT2 inhibitor use were accounted for in the mixed-effects models, other dopaminergic medications (such as carbidopa-levodopa) were not factored into the analysis. The use of DTW, while mathematically robust and appropriate for time series analyses, is not associated with a direct p-value to assess for statistical significance.(32) Although statistical methods were employed to complement this lack of p-value, it nevertheless remains a drawback of using DTW. Furthermore, the claim that similarity in subscale trajectory implies similarity in pathoetiology is an assumption and may be proven wrong considering objective biological evidence. Despite these shortcomings, the conclusions derived from this paper allow us to classify the 31 UHDRS motor subscales with some degree of confidence. More importantly, perhaps, it allows for more questions to be posed in light of these findings, such as the effect of VMAT2 inhibitors on both hyperkinetic and hypokinetic features alike.

## Data Availability

The datasets utilized for this secondary analysis are obtainable from the Enroll-HD clinical research platform, courtesy of CHDI, for use by professional researchers in their studies on HD.

https://enroll-hd.org/for-researchers/access-data-biosamples/

## Acknowledgements

This research is made possible due to the efforts of CHDI, the *Enroll-HD* clinical research platform and the participation of HD patients and their families.

## Authors’ Roles

Research Project: A. Conception, B. Organization, C. Execution; 2. Statistical Analysis: A. Design, B. Execution, C. Review and Critique; 3. Manuscript Preparation: A. Writing the First Draft, B. Review and Critique.

N.M.H.: 1A, 1B, 1C, 2A, 2B, 3A, 3B

J.L.S.: 1A, 1B, 1C, 2A, 2B, 3A, 3B P.C.N.: 3B

A.K.: 3B

## Disclosures

**Ethical Compliance Statement**: The *Enroll-HD* clinical platform, funded and run by CHDI (Cure Huntington’s Disease Foundation), has conducted their original study under the oversight and approval of multiple Institutional Review Boards. More specifically, each *Enroll-HD* and *Registry* clinical site has their own consent form and consent-obtaining process, overseen by the local IRB at their institution. Part of this consent involves informing participants that their *de-identified* data may be made available to qualified researchers at a later date.

**Funding Sources and Conflict of Interest**: Jordan L. Schultz was supported by the National Institute of Neurological Disability and Stroke (NINDS) through the following grant: K23-NS117736. Jordan L. Shultz is a paid consultant of the Huntington Study Group (HSG). Dr. Nopoulos was supported by the NINDS through the following grant: U01NS055903-13. Dr. Nopoulos was supported by the National Institute of Child Health and Human Development (NICHD) through the following grant: P50HD103556-04. Dr. Nopoulos serves as a member of the scientific advisory board for uniQure. The authors declare that there are no conflicts of interest relevant to this work.

**Financial disclosures from previous 12 months**: Enroll-HD is a multi-site longitudinal study which follows the natural history of HD patients and their non-affected relatives; it is sponsored in its entirety by the CHDI, a not-for-profit research organization which dedicates its funds and research to HD.

