## Supplement for "Differentiating Hyperkinetic and Hypokinetic Motor Features in the Progression of Huntington’s Disease"

*Supplemental Table 1: Significance of Disease Burden with Respect to Motor Subscale Progression*

| **Subscale** | **Sum of Squares** | **Mean Squared** | **Numerator Degrees of Freedom** | **Denominator Degrees of Freedom** | **F-Value** | **P-Value** |
| --- | --- | --- | --- | --- | --- | --- |
| **Bradykinesia** | 4028.462 | 1342.821 | 3 | 26497.24 | 2705.084 | <0.0001 |
| **Retropulsion** | 3313.677 | 1104.559 | 3 | 24369.61 | 2240.554 | <0.0001 |
| **Rigidity Arm Left** | 1672.894 | 557.631 | 3 | 24226.67 | 1521.974 | <0.0001 |
| **Rigidity Arm Right** | 1619.650 | 539.883 | 3 | 24193.63 | 1491.962 | <0.0001 |
| **Chorea BOL** | 895.240 | 298.413 | 3 | 22387.76 | 599.254 | <0.0001 |
| **Chorea Face** | 693.147 | 231.049 | 3 | 22898.83 | 521.577 | <0.0001 |
| **Chorea Trunk** | 739.512 | 246.504 | 3 | 22747.70 | 469.033 | <0.0001 |
| **Chorea RUE** | 580.929 | 193.643 | 3 | 23294.87 | 434.997 | <0.0001 |
| **Chorea LUE** | 573.424 | 191.141 | 3 | 23349.98 | 428.854 | <0.0001 |
| **Chorea RLE** | 323.664 | 107.888 | 3 | 22863.50 | 232.997 | <0.0001 |
| **Chorea LLE** | 322.471 | 107.490 | 3 | 22830.30 | 230.444 | <0.0001 |

Abbreviations: LUE, left-upper extremity; RUE, right-upper extremity; LLE, left-lower extremity; RLE, right-lower extremity; BOL, buccal-oral-lingual; H, horizontal; V, vertical.

*Supplementary Table 2: Summary of Simplistic DTW Motor Subscale Classification*

| **Subscale** | **Classification** | **Distance to Hyperkinetic Curve** | **Distance to Hypokinetic Curve** |
| --- | --- | --- | --- |
| **Ocular H.** | Hypokinetic | 447.056034 | 26.68983 |
| **Ocular V.** | Hypokinetic | 547.756346 | 67.81866 |
| **Saccadic Initiation H.** | Hypokinetic | 610.655829 | 66.56368 |
| **Saccadic Initiation V.** | Hypokinetic | 649.414707 | 84.03509 |
| **Saccadic Velocity H.** | Hypokinetic | 630.640297 | 69.38832 |
| **Saccadic Velocity V.** | Hypokinetic | 702.293916 | 119.07975 |
| **Dysarthria** | Hypokinetic | 490.044382 | 54.60748 |
| **Tongue Protrusion** | Hypokinetic | 544.584872 | 34.68338 |
| **Finger Tapping Right** | Hypokinetic | 698.076151 | 90.56133 |
| **Finger Tapping Left** | Hypokinetic | 770.526769 | 132.52645 |
| **Pronation Supination Right** | Hypokinetic | 677.140262 | 70.14865 |
| **Pronation Supination Left** | Hypokinetic | 722.845734 | 109.49542 |
| **Luria** | Hypokinetic | 713.256570 | 109.93508 |
| **Rigidity Right Arm** | Hypokinetic | 257.124988 | 44.21870 |
| **Rigidity Left Arm** | Hypokinetic | 265.225951 | 44.71635 |
| **Bradykinesia Body** | Hypokinetic | 640.881048 | 60.23415 |
| **Truncal Dystonia** | Hypokinetic | 315.047255 | 79.04009 |
| **RUE Dystonia** | Hypokinetic | 315.761775 | 88.25189 |
| **LUE Dystonia** | Hypokinetic | 315.051811 | 91.87008 |
| **RLE Dystonia** | Hypokinetic | 230.289260 | 177.89384 |
| **LLE Dystonia** | Hypokinetic | 231.229627 | 181.17833 |
| **Facial Chorea** | Hyperkinetic | 6.665345 | 339.17686 |
| **BOL Chorea** | Hyperkinetic | 11.524983 | 336.09804 |
| **Truncal Chorea** | Hyperkinetic | 8.099901 | 377.11922 |
| **RUE Chorea** | Hyperkinetic | 18.068329 | 396.94683 |
| **LUE Chorea** | Hyperkinetic | 17.286443 | 391.81662 |
| **RLE Chorea** | Hyperkinetic | 39.405185 | 492.50743 |
| **LLE Chorea** | Hyperkinetic | 37.913379 | 481.09137 |
| **Gait** | Hypokinetic | 599.574546 | 55.65500 |
| **Tandem-Walking** | Hypokinetic | 937.617658 | 234.62646 |
| **Retropulsion Test** | Hypokinetic | 490.066054 | 23.80118 |

Abbreviations: DTW, dynamic time warping; LUE, left-upper extremity; RUE, right-upper extremity; LLE, left-lower extremity; RLE, right-lower extremity; BOL, buccal-oral-lingual; H, horizontal; V, vertical.

*Supplementary Table 3: Magnitudes and P-values of Antipsychotic-Burden and VMAT2 inhibitor-Burden Interaction Term Coefficients of the Motor Subscale Linear Models*

| **Subscale** | **Coefficient** | **Estimate** | **Standard Error** | **P-value** |
| --- | --- | --- | --- | --- |
| **Ocular Pursuit Horizontal** | Antipsychotic-Burden Interaction | 0.318406501 | 0.064586103 | 8.29 X 10-07 |
| **Ocular Pursuit Horizontal** | VMAT2i-Burden Interaction | 0.396912659 | 0.07784859 | 3.45 X 10-07 |
| **Ocular Pursuit Vertical** | Antipsychotic-Burden Interaction | 0.453310762 | 0.066403758 | 8.93 X 10-12 |
| **Ocular Pursuit Vertical** | VMAT2i-Burden Interaction | 0.403280394 | 0.080036356 | 4.73 X 10-07 |
| **Saccade Initiation Horizontal** | Antipsychotic-Burden Interaction | 0.275077404 | 0.06551996 | 2.70 X 10-05 |
| **Saccade Initiation Horizontal** | VMAT2i-Burden Interaction | 0.128822873 | 0.078979444 | 1.03 X 10-01 |
| **Saccade Initiation Vertical** | Antipsychotic-Burden Interaction | 0.360125573 | 0.06600613 | 4.92 X 10-08 |
| **Saccade Initiation Vertical** | VMAT2i-Burden Interaction | 0.188855581 | 0.079571807 | 1.76 X 10-02 |
| **Saccade Velocity Horizontal** | Antipsychotic-Burden Interaction | 0.351407243 | 0.065841717 | 9.54 X 10-08 |
| **Saccade Velocity Horizontal** | VMAT2i-Burden Interaction | 0.320093126 | 0.079362295 | 5.52 X 10-05 |
| **Saccade Velocity Vertical** | Antipsychotic-Burden Interaction | 0.394425871 | 0.069615204 | 1.48 X 10-08 |
| **Saccade Velocity Vertical** | VMAT2i-Burden Interaction | 0.366759588 | 0.083899175 | 1.24 X 10-05 |
| **Dysarthria** | Antipsychotic-Burden Interaction | 0.48863916 | 0.054962609 | 6.56 X 10-19 |
| **Dysarthria** | VMAT2i-Burden Interaction | 0.415321944 | 0.066272881 | 3.75 X 10-10 |
| **Tongue Protrusion** | Antipsychotic-Burden Interaction | 0.611100127 | 0.068739636 | 6.57 X 10-19 |
| **Tongue Protrusion** | VMAT2i-Burden Interaction | 0.148937809 | 0.082868821 | 7.23 X 10-02 |
| **Finger-Tapping Right** | Antipsychotic-Burden Interaction | 0.497375006 | 0.063551514 | 5.25 X 10-15 |
| **Finger-Tapping Right** | VMAT2i-Burden Interaction | 0.274735566 | 0.076626478 | 3.37 X 10-04 |
| **Finger-Tapping Left** | Antipsychotic-Burden Interaction | 0.444227589 | 0.062053212 | 8.40 X 10-13 |
| **Finger-Tapping Left** | VMAT2i-Burden Interaction | 0.197778869 | 0.074817839 | 8.21 X 10-03 |
| **Pro/Supination Right Hand** | Antipsychotic-Burden Interaction | 0.622154162 | 0.066912504 | 1.56 X 10-20 |
| **Pro/Supination Right Hand** | VMAT2i-Burden Interaction | 0.324216683 | 0.080681384 | 5.88 X 10-05 |
| **Pro/Supination Left Hand** | Antipsychotic-Burden Interaction | 0.564718454 | 0.065506103 | 7.09 X 10-18 |
| **Pro/Supination Left Hand** | VMAT2i-Burden Interaction | 0.232846874 | 0.078985451 | 3.20 X 10-03 |
| **Luria** | Antipsychotic-Burden Interaction | 0.446995273 | 0.088018002 | 3.84 X 10-07 |
| **Luria** | VMAT2i-Burden Interaction | 0.131453475 | 0.106104215 | 2.15 X 10-01 |
| **Rigidity Right Arm** | Antipsychotic-Burden Interaction | 0.119136772 | 0.053837684 | 2.69 X 10-02 |
| **Rigidity Right Arm** | VMAT2i-Burden Interaction | 0.167947803 | 0.064822589 | 9.58 X 10-03 |
| **Rigidity Left Arm** | Antipsychotic-Burden Interaction | 0.121059157 | 0.054382767 | 2.60 X 10-02 |
| **Rigidity Left Arm** | VMAT2i-Burden Interaction | 0.136842672 | 0.06548404 | 3.67 X 10-02 |
| **Bradykinesia - Body** | Antipsychotic-Burden Interaction | 0.47457604 | 0.071034546 | 2.43 X 10-11 |
| **Bradykinesia - Body** | VMAT2i-Burden Interaction | 0.237199335 | 0.085651336 | 5.62 X 10-03 |
| **Dystonia Trunk** | Antipsychotic-Burden Interaction | 0.368922294 | 0.063415793 | 6.06 X 10-09 |
| **Dystonia Trunk** | VMAT2i-Burden Interaction | 0.206387752 | 0.07631511 | 6.85 X 10-03 |
| **Dystonia Right Upper Extremity** | Antipsychotic-Burden Interaction | 0.262742001 | 0.058739598 | 7.75 X 10-06 |
| **Dystonia Right Upper Extremity** | VMAT2i-Burden Interaction | 0.113186243 | 0.070649023 | 1.09 X 10-01 |
| **Dystonia Left Upper Extremity** | Antipsychotic-Burden Interaction | 0.255523549 | 0.058596406 | 1.30 X 10-05 |
| **Dystonia Left Upper Extremity** | VMAT2i-Burden Interaction | 0.130866873 | 0.070482693 | 6.34 X 10-02 |
| **Dystonia Right Lower Extremity** | Antipsychotic-Burden Interaction | 0.219167739 | 0.055025428 | 6.83 X 10-05 |
| **Dystonia Right Lower Extremity** | VMAT2i-Burden Interaction | 0.133729101 | 0.066163436 | 4.33 X 10-02 |
| **Dystonia Left Lower Extremity** | Antipsychotic-Burden Interaction | 0.236348875 | 0.054436731 | 1.42 X 10-05 |
| **Dystonia Left Lower Extremity** | VMAT2i-Burden Interaction | 0.139897035 | 0.065454647 | 3.26 X 10-02 |
| **Facial Chorea** | Antipsychotic-Burden Interaction | -0.006492429 | 0.056038218 | 9.08 X 10-01 |
| **Facial Chorea** | VMAT2i-Burden Interaction | -0.192873633 | 0.067366204 | 4.20 X 10-03 |
| **Buccal/Oral/Lingual Chorea** | Antipsychotic-Burden Interaction | -0.011587514 | 0.058787941 | 8.44 X 10-01 |
| **Buccal/Oral/Lingual Chorea** | VMAT2i-Burden Interaction | -0.140145878 | 0.070649145 | 4.73 X 10-02 |
| **Truncal Chorea** | Antipsychotic-Burden Interaction | -0.001981791 | 0.060643764 | 9.74 X 10-01 |
| **Truncal Chorea** | VMAT2i-Burden Interaction | -0.203819168 | 0.072888726 | 5.17 X 10-03 |
| **Chorea Right Upper Extremity** | Antipsychotic-Burden Interaction | -0.203128244 | 0.056752149 | 3.45 X 10-04 |
| **Chorea Right Upper Extremity** | VMAT2i-Burden Interaction | -0.378119802 | 0.068244701 | 3.05 X 10-08 |
| **Chorea Left Upper Extremity** | Antipsychotic-Burden Interaction | -0.203679451 | 0.056855741 | 3.41 X 10-04 |
| **Chorea Left Upper Extremity** | VMAT2i-Burden Interaction | -0.359610027 | 0.068371578 | 1.46 X 10-07 |
| **Chorea Right Lower Extremity** | Antipsychotic-Burden Interaction | -0.075056415 | 0.057076223 | 1.89 X 10-01 |
| **Chorea Right Lower Extremity** | VMAT2i-Burden Interaction | -0.281368713 | 0.06860623 | 4.13 X 10-05 |
| **Chorea Left Lower Extremity** | Antipsychotic-Burden Interaction | -0.079182591 | 0.057206345 | 1.66 X 10-01 |
| **Chorea Left Lower Extremity** | VMAT2i-Burden Interaction | -0.263760804 | 0.068759721 | 1.25 X 10-04 |
| **Gait** | Antipsychotic-Burden Interaction | 0.498540383 | 0.060033689 | 1.06 X 10-16 |
| **Gait** | VMAT2i-Burden Interaction | 0.539053912 | 0.072385745 | 9.92 X 10-14 |
| **Tandem-Walking** | Antipsychotic-Burden Interaction | 0.718106917 | 0.080660763 | 5.87 X 10-19 |
| **Tandem-Walking** | VMAT2i-Burden Interaction | 0.344082847 | 0.097249121 | 4.04 X 10-04 |
| **Retropulsion Pull Test** | Antipsychotic-Burden Interaction | 0.520449816 | 0.066842261 | 7.22 X 10-15 |
| **Retropulsion Pull Test** | VMAT2i-Burden Interaction | 0.438925707 | 0.080566643 | 5.15 X 10-08 |

Abbreviations: VMAT2, vesicular monoamine transporter type 2.
